# Research priorities for improving menstrual health across the life-course in low- and middle-income countries

**DOI:** 10.1101/2023.01.31.23285290

**Authors:** Marina Plesons, Belen Torondel, Bethany A. Caruso, Julie Hennegan, Marni Sommer, Jacqueline Haver, Danielle Keiser, Anna M van Eijk, Garazi Zulaika, Linda Mason, Penelope A. Phillips-Howard

## Abstract

**Background:** Research on menstrual health is required to understand the needs of girls, women, and others who menstruate; and to strengthen programmes with evidence-based interventions to improve health, wellbeing, and productivity. The identification of research priorities is an important process to help researchers, policymakers, programmers, and funding agencies decide where to invest their efforts and resources.

**Methods:** A modified version of the Child Health and Nutrition Research Initiative (CHNRI) approach was utilized to reach consensus on a set of research priority questions. Multisector stakeholders with expertise in policy, programming, financial support, and/or research relating to menstrual health were identified through networks and the literature. Individuals were invited to submit priority questions through survey monkey online on i) understanding the problem; ii) designing and implementing interventions; iii) integrating and scaling up interventions. Responses were consolidated and individuals were then invited to rank these questions based on i) novelty; ii) potential for intervention; and iii) importance/impact. Research priority scores were calculated from these responses and analyzed to evaluate associations with social and occupational characteristics of participating respondents.

**Results:** Eighty-two participants, of whom 89% were female and 39.0% from low- and middle-income countries (LMIC), proposed a total of 1135 research questions: 45.9% on understanding the problem, 39.7% on designing and implementing interventions, 12.7% on integrating and scale-up, plus 1.5% on other aspects. Questions were consolidated into a final list of 91 unique research questions. Sixty-six participants, of whom 80.3% were women and 39.4% from LMIC, ranked these questions. Top ten-ranked research priority questions comprised four questions on ‘understanding the problem’, four on ‘designing and implementing interventions’, one on ‘integrating and scaling up’, and one on ‘measurement and research’. Academics gave higher prioritization to ‘designing and implementing interventions,’ and lower prioritization to questions on ‘understanding the problem,’ ‘integrating and scaling up,’ and ‘measurement and research.

**Conclusions:** Use of CHNRI generated unique research priority questions from expertise internationally. The top-ranking research priorities can be utilized by policymakers, programmers, researchers, and funders to guide future research in menstrual health.

## Introduction

Menstrual health has received increased attention in recent years as an important component of public health.^1,2^ Research in low- and middle-income countries (LMICs) – largely descriptive studies – has identified the impacts of poor menstrual health on girls’ health, wellbeing, and education.^3,4^ A small number of recent trials have evaluated the impact of menstrual products and puberty education on girls’ school attendance, educational performance, sexual and reproductive health (SRH), and wellbeing.^5–8^ Other studies have focused on understanding the menstrual self-care practices and menstrual health challenges of women and girls in humanitarian contexts, as well as the acceptability of specific menstrual products in such settings.^9–11^ More recently, studies have started to describe challenges for adult women,^12–15^marginalized populations in high income countries (HICs),^16–20^, mental health,^21^ and to develop measures for menstrual health research.^22,23^ The evidence on menstrual health has been consolidated in a growing body of systematic reviews, including reviews focused on specific geographies,^24–26^ populations (e.g. girls with disabilities,^27^ those who are displaced),^24,28^ measures of exposures and outcomes,^29 23^ interventions (e.g. menstrual cups, reusable menstrual pads),^30^{van Eijk, 2021 #3976} and outcomes (e.g. knowledge and understanding, health, and social wellbeing).^4,26,31–33^

The identification of research priorities is an important process to help researchers, programmers, policymakers, and funding agencies decide where to invest their efforts and resources. In 2014, research priorities on menstrual hygiene management (MHM) among school-going girls in LMICs were identified by an expert group as part of “MHM in Ten,” an initiative that sought to set the agenda for overcoming challenges related to menstrual health faced by this population and for identifying the evidence needed to improve girls’ experiences of menstruation and education.^1,3^ As evidenced by its extensive citation and the substantial number of relevant research outputs since 2014, it is clear that the prioritization effort has positively impacted the trajectory of the field, which has developed rapidly in the eight years since. Several identified priorities have been acted upon, including the need for strengthening the evidence base. A review of qualitative studies on menstrual health reported that 50% of the studies were published between 2015 and 2019.^4^ Other priorities have also seen progress, including the need for standardized menstrual measures^34^ and definitions,^23,35,36^, and the need for a research consortium, which has begun to take shape under the umbrella of the Global Menstrual Collective. Given the evolution of the field, it is therefore timely to reassess the research priorities for improving menstrual health to guide the field. Further, as the focus of efforts to address menstrual health has expanded beyond school-going girls to include girls who are out-of-school, as well as women and others who menstruate, there is a need to identify research priorities to address the needs of all who menstruate across the life-course in varying contexts around the world.^37^

Thus, the objective of this study is to identify research priorities for menstrual health across the life-course in LMICs, in consultation with a range of stakeholder groups from a variety of geographic regions. This study additionally aims to understand if and how research priorities differ across sectors, stakeholder groups, and years working in menstrual health.

## Methods

This research was undertaken by members of the Global Menstrual Collective’s Research and Evidence Group, comprising researchers, stakeholders, and implementers with experience working in the field of menstrual health. The Global Menstrual Collective is a collaborative network whose aim is to bring together partners in menstrual health to amplify efforts and reduce duplication for mainstreaming menstrual health across health, education, gender, and water, sanitation, and hygiene (WASH).

We used a modified version of the Child Health and Nutrition Research Initiative (CHNRI) approach to reach consensus on a set of research priority questions. The CHNRI approach is a transparent and structured process for ranking the relative importance of competing research priorities to help decision-makers effectively allocate limited resources to address a health problem, e.g. by reducing morbidity and mortality, improving wellbeing and quality of life, and addressing inequities.^38,39^ It has been used to reach consensus on research priorities for numerous health topics, including adolescent health,^40^ adolescent sexual and reproductive health,^41^ and family planning.^42^ The adapted CHINRI approach used in this study involved three phases, described in detail below:

### Phase 1: Identification of individuals with expertise in menstrual health

First, individuals from various stakeholder groups with expertise in policy, programming, financial support, and/or research relating to menstrual health were identified. This was achieved by disseminating information about the study to (i) menstrual health panels, coalitions, and consortia (e.g., the Menstrual Health Hub, the African Coalition for Menstrual Health Management, the MHM in Ten Expert network, Water Supply and Sanitation Collaborative Council and the GMC Collective, the Menstrual Cup Collaboration, and Water Sanitation and Hygiene (WASH) United’s Menstrual Hygiene Day), (ii) published researchers, and (iii) funders identified through past or current funding calls.

Snowballing was then used to circulate the invitation to relevant others who may have been missed by the above methods.

### Phase 2: Identification of research questions on menstrual health

The individuals identified in Phase 1 were invited to propose research questions on menstrual health across the life-course, using an electronic survey (SurveyMonkey, Palo Alto). An invitation email was sent in September 2020 with two reminders sent fortnightly. The survey was closed in October 2020.

After reading an information sheet explaining the nature and purpose of the exercise, consenting to participate, and providing demographic information including sector of work (e.g., academia, UN-agency, non-governmental organization) and geographic areas of residence and work, participants were prompted to propose research questions. These spanned three domains (each with several sub-domains), as guided by the CHNRI approach:

1. *Understanding the problem*: questions to illustrate the experiences of those who menstruate, explore risk and protective factors for menstrual health, and test impacts and consequences of poor menstrual health. Such questions could utilize a range of methodologies, from descriptive epidemiology to ethnographic research.
2. *Designing and implementing interventions*: questions which relate to (i) discovery of new interventions, (ii) development and testing of interventions/programmes, and (iii) evaluations of the effectiveness, acceptability, adoption, appropriateness, feasibility, fidelity, cost, coverage, and sustainability of interventions to address menstrual health. Such questions could utilize intervention effectiveness research and implementation research.
3. *Integrating and scaling up interventions*: questions which relate to integrating menstrual health interventions into health, education, WASH, or social services and to taking menstrual health interventions to scale. Such questions could include implementation research and policy and systems research.

Exemplar questions for each domain and sub-domain were provided to provide further clarity for the participants.

Participant responses were downloaded into spreadsheets, and free texts were collapsed to aggregate group data. A core team of four members of the Global Menstrual Collective’s Research and Evidence Group then iteratively categorized and consolidated the questions based on themes (Tables S1). Further bracketing of questions between domains and sub-domains was undertaken where relevant. Duplicates were removed, as were questions covering unrelated topics. An extra domain, *Measurement & Research*, was included as numerous questions pertaining to this topic were suggested by participants. Similar questions were condensed together to derive a smaller number of amalgamated research questions (Table S1). Once the full set of consolidated questions was developed, a meeting with the Global Menstrual Collective’s Research and Evidence Group was held to review and agree upon a final list of research questions to be used in Phase 3.

### Phase 3: Prioritization of the proposed research questions on menstrual health

Discussion among the Global Menstrual Collective’s Research and Evidence Group raised concerns about the length of the survey, the clarity of these criteria for this specific topic, and potential confounding due to explicit mention of equity in multiple questions. To address these concerns, the five criteria originally proposed by the CHNRI approach (clarity, answerability, importance/impact, implementation, equity) were modified. The modifications were in line with the CHNRI approach, which suggests that the priority setting process should list possible criteria appropriate to their specific context and may merge criteria, where appropriate.^39^ Thus, three criteria – novelty, potential for implementation, and importance/impact (Table 1) – were agreed upon, with the CHNRI approach’s standard scoring system of *yes, no, or undecided*. Due to its length, the survey was split into two sections comprising 40 and 51 research questions.

**Table 1.** Criteria used to score the proposed research questions and their definitions.

|  | Understanding the problem | Designing and implementing interventions | Integrating and scaling-up interventions | Measurement & research |
| --- | --- | --- | --- | --- |
| **Novelty** | Will the answer to this question fill a key gap? |  |  |  |
| **Potential for implementation** | Will the answer to the question contribute to tailored interventions? | Will the answer to the question result in an intervention that can be implemented? | Could the answer to the question be implemented? |  |
| **Importance/impact** | Will the answer to this question be important to know? | Will the answer to the question result in an intervention that would have an important impact? | Will the answer to the question have an important impact? |  |

The individuals identified in Phase 1 were then invited to score the proposed research questions, again using an electronic survey (SurveyMonkey, Palo Alto). An invitation email was sent in June 2021 with two reminders sent fortnightly. The survey was closed in July 2021.

Participant responses were downloaded into spreadsheets, cleaned, and imported into IBM SPSS version 28 (Armonk). A variable was created to indicate if participants had responded to individual research priority questions. The number of responses per participant and per research question were counted.

For participant demographic information, frequency distributions of characteristics of the participants were conducted. For ranking of the research questions, a score of 100 points was attributed to “yes”, 50 points to “undecided”, and 0 points to “no”. A total Research Priority Score (RPS) was then assigned for each research question by computing the mean score across the three criteria. RPSs were then ranked from highest to lowest, overall and within each domain. RPSs were also assessed based on the profile of the participants, e.g., by their sector of work, stakeholder group, and years working in menstrual health.

### Ethical Considerations

The project was approved by the Liverpool School of Tropical Medicine’s Research and Ethics Committee (ID# 20-055), as exemption given by the Human Reproduction Programme Research Protocol Review Panel and the WHO Ethics Review Committee (ID# ERC.0003407). Potential respondents were informed that their participation was voluntary, and they were free to stop responding to questions at any time.

Written consent was obtained through the provision of a participant information sheet and consent on the online webpage. Participants had to tick that they consented to join the study before they could access the survey.

## Results

### Characteristics of Phase 2 participants

A total of 82 participants responded to the Phase 2 survey and proposed research questions on menstrual health (Table 2). The majority were female (89%) with 33% and 28% aged 25-34 years and 35-44 years respectively. The highest proportion (61%) originated from HICs, with 29% from Europe; LMICs were represented, with 27% of participants from sub-Saharan Africa. The highest proportion of respondents worked in non-government organization (NGO) and international NGO (43%) or in academia (35%). A third of participants worked globally, over half (58%) worked in sub-Saharan Africa, and 35% worked in east and southern Asia. Sixty-two percent of participants reported their area of expertise lay in sexual and reproductive health (SRH). Over one third (38%) of participants had worked in the field of menstrual health for 7 years or longer.

**Table 2.**
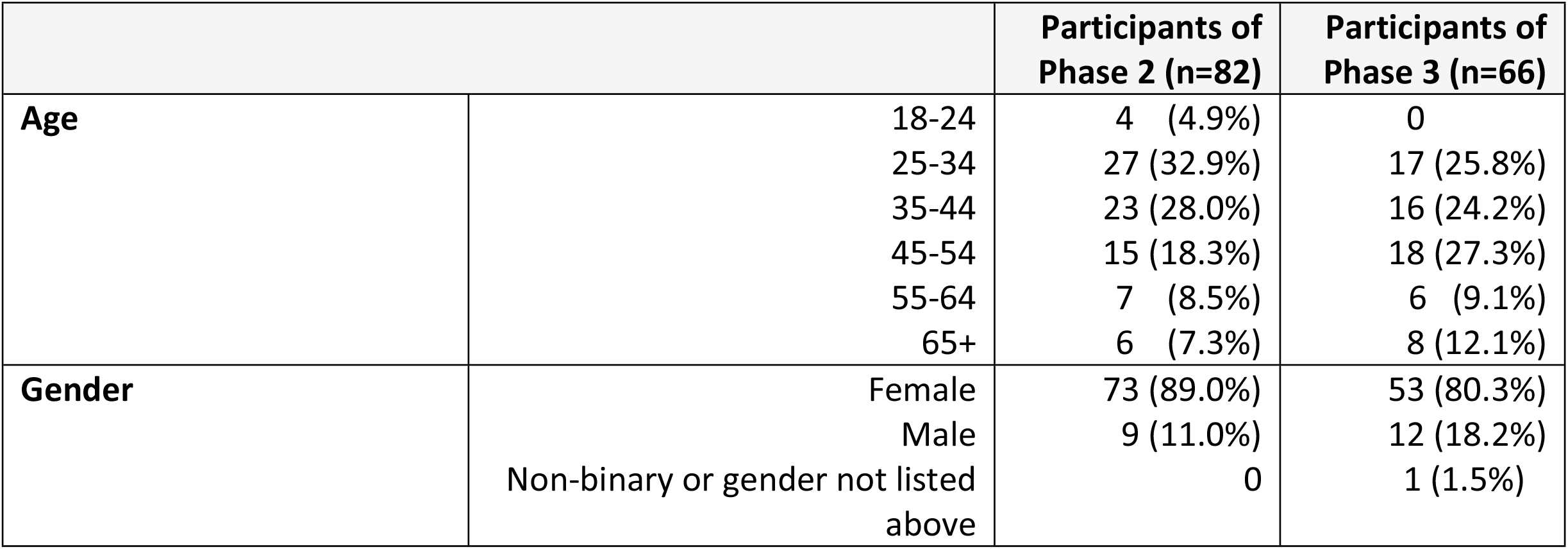
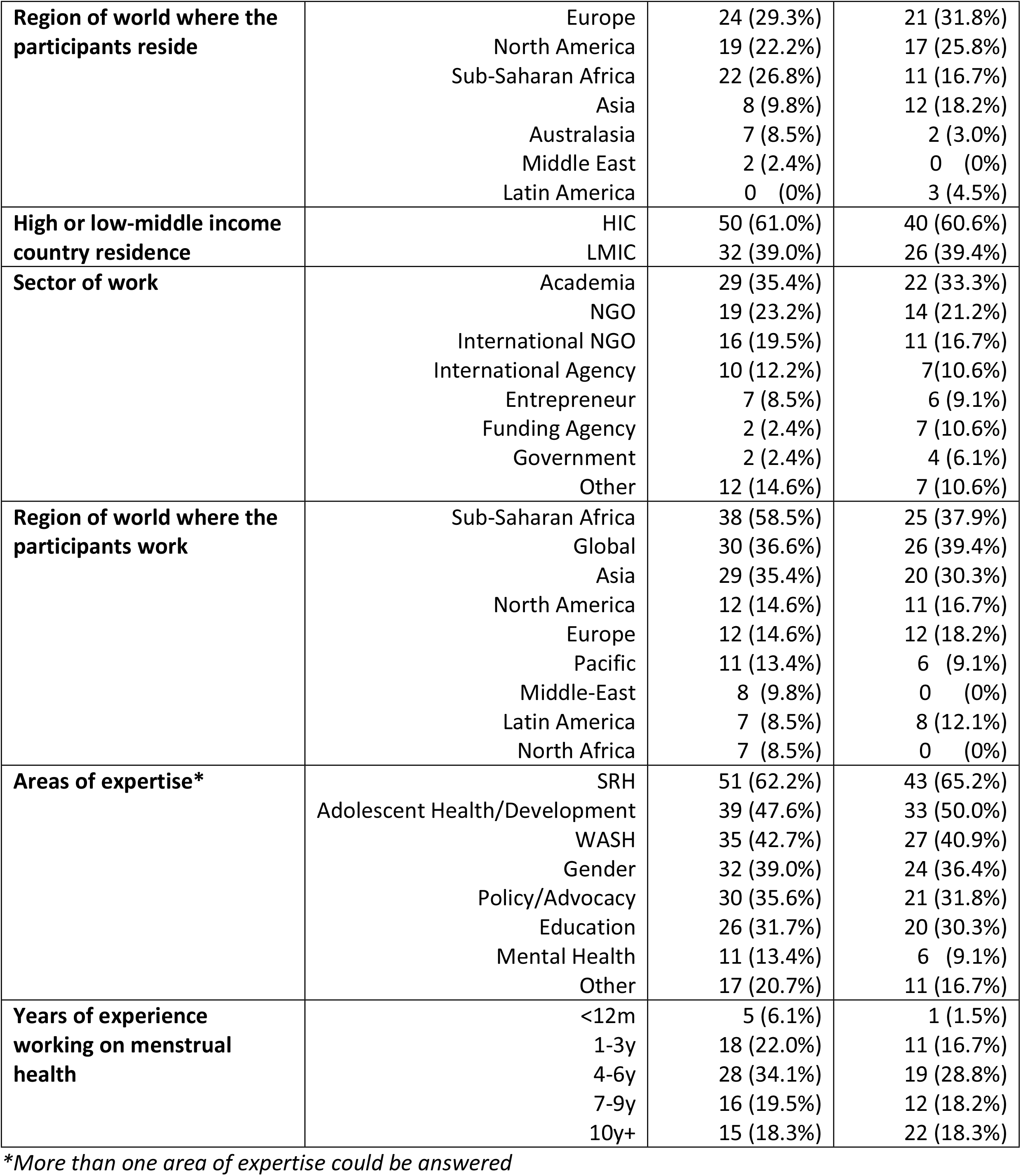
Participant characteristics (n=82)

### Proposed research questions

A total of 1135 research questions were proposed by the 82 participants that responded to the Phase 2 survey, with an average of 13.8 (standard deviation [sd 9.9]; median 11) research questions proposed per participant (Table 3 and Table S2). The greatest number of research questions proposed were on understanding the problem (521 by all participants; average 6.4 per participant), followed by designing and implementing interventions (451 by 69 participants, average 6.5 per participant). Participants with more years of experience in menstrual health (7 years or more) proposed considerably more research questions (average 17.6), compared with those with fewer years of experience (average 11.7) (Table S3). Participants working in the mental health sector proposed significantly more research questions (average 19.7) than those working in other areas (combined average 13.17). The differences among stakeholder groups were less pronounced.

**Table 3.** Overview of the proposed research questions.

| Domain | Total number of research questions proposed | Number of participants that proposed at least one research question | Average number of research questions proposed per participant |
| --- | --- | --- | --- |
| Understanding the problem | 521 (45.9%) | 82 | 6.4 |
| Designing and implementing interventions | 451 (39.7%) | 69 | 6.5 |
| Integrating and scale-up | 145 (12.7%) | 54 | 2.7 |
| Other | 18 (1.6%) | 15 | 1.2 |
| **Total** | **1135** |  | **13.8** |

As previously described, the proposed research questions were consolidated into a final list of 91 unique research questions. A breakdown of the number of research questions per domain and sub-domain is provided in Table 4.

**Table 4.** Overview of the consolidated list of research questions.

| Domain | Sub-domain | Number of research questions |
| --- | --- | --- |
| Understanding the problem | Experiences related to menstrual health | 7 |
|  | Factors affecting menstrual health | 9 |
|  | Impact and consequences | 7 |
|  | **Total** | **23** |
| Designing and implementing interventions | Discovery of new interventions | 12 |
|  | Developing and testing the effectiveness of interventions/programmes | 18 |
|  | Evaluations of the costs of interventions/programmes | 6 |
|  | Evaluations of the delivery (including acceptability, adoption, appropriateness, feasibility, fidelity, coverage, and reach) of interventions/ programmes | 10 |
|  | Evaluations of the sustainability of interventions/programmes | 6 |
|  | **Total** | **52** |
| Integrating and scale-up | Integration | 6 |
|  | Scale-up | 5 |
|  | **Total** | **11** |
| Measurement & Research |  | **5** |
| **Grand Total** |  | **91** |

### Characteristics of Phase 3 participants

A total of 66 participants completed the Phase 3 survey, contributing to the prioritization of the proposed research questions (Table 2). The majority were female (80%), with 26% and 27% aged 25-34 years and 45-54 years respectively. The highest proportion (61%) resided in HICs, with 32% of the participants from Europe; LMICs were represented by 39%, with 18% of the participants from Asia. The highest proportion of respondents worked in NGO or international NGO (38%) and academia (33%) sectors. Over a third (39%) of participants worked globally, 38% worked in Sub-Saharan Africa, and 30% worked in east and southern Asia. Sixty-five percent of participants reported working in SRH. Over one third (36%) had worked in the field of menstrual health for 7 years or more.

### Prioritized research questions

The highest ranked research question by RPS among all participants was ‘What indicators are optimal for assessing menstrual health over time (e.g., related to norms, education, health, rights, etc.)?’ It was ranked highest according to non-academic participants, and second highest according to academic participants.

The top ten-ranked research questions are listed in Table 5. They include four questions on ‘understanding the problem’, four on ‘designing and implementing interventions’, one on ‘integrating and scaling up’, and one on ‘measurement and research’. We found a high level of agreement on these ten questions, with total RPS ranging from 0.913 to 0.956 (out of a possible 1). Of note, the difference in RPS between the top ten-ranked research questions and those subsequent was not substantial, pointing to the high prioritization of many questions beyond those included only in this abbreviated list (Figure 1). Thus, the top five-ranked research questions in each domain, ranked according to their RPS, are listed in Table 6.

**Table 5.** Top 10-ranked research questions, by Research Priority Score.

| Rank | RPS | Question | Domain | Unique ID |
| --- | --- | --- | --- | --- |
| 1 | 0.956 | What indicators are optimal for assessing menstrual health over time (e.g. related to norms, education, health, rights, etc.)? | Measurement and research | M1 |
| 2 | 0.932 | What are the experiences of girls, women, and others who menstruate in relation to menstrual pain and disorders (e.g. what proportion experience them, what are their perceptions about them, how do they manage them, and what support do they seek and receive for them)? | Understanding the problem | U1 |
| 3 | 0.93 | What new interventions could be developed to address harmful attitudes, norms and stigma and improve communication related to menstruation? | Designing and implementing interventions | D1 |
| 4 | 0.924 | Are girls, women, and others who menstruate able to access and afford their preferred menstrual product/materials; what is the quality of these products/materials; where do they obtain them; and how do they use and dispose of them? | Understanding the problem | U2 |
| 5 | 0.921 | What is the impact of unmet menstrual health needs (e.g. products/materials, WASH infrastructure/services) on the participation and engagement of girls, women, and others who menstruate in school and work and their self-esteem and agency? | Understanding the problem | U3 |
| 6 | 0.919 | What are the experiences and challenges of girls, women, and others who menstruate with particular needs and circumstances (e.g. those living with HIV, those with disabilities, those who are incarcerated, those experiencing homelessness, those who have experienced FGM, trans and gender non-binary persons) in relation to their menstrual health? | Understanding the problem | U4 |
| 7 | 0.915 | What characteristics of menstrual health interventions/programmes enable them to be sustained over time? | Designing and implementing interventions | D2 |
| 8 | 0.915 | How can interventions to address menstrual health (e.g. education, social norm change, distribution of menstrual products/materials, improvements in WASH infrastructure/services, provision of health services for menstrual pain and disorders) be scaled up with quality and equity? | Integrating and scaling up | I1 |
| 9 | 0.913 | What new interventions could be developed to manage menstrual pain? | Designing and implementing interventions | D3 |
| 10 | 0.913 | What are the impacts of unconditional and conditional cash transfer interventions on the menstrual health of girls, women, and others who menstruate, and consequently on their education, work, and social participation? | Designing and implementing interventions | D4 |

**Table 6.** Top five-ranked research questions in each domain, by Research Priority Score.

| Domain | Rank within domain | Overall rank | RPS | Question | Unique ID |
| --- | --- | --- | --- | --- | --- |
| **Understanding the problem (n=25)** | 1 | 2 | 0.932 | What are the experiences of girls, women, and others who menstruate in relation to menstrual pain and disorders (e.g. what proportion experience them, what are their perceptions about them, how do they manage them, and what support do they seek and receive for them)? | U1 |
|  | 2 | 4 | 0.924 | Are girls, women, and others who menstruate able to access and afford their preferred menstrual product/materials; what is the quality of these products/materials; where do they obtain them; and how do they use and dispose of them? | U2 |
|  | 3 | 5 | 0.921 | What is the impact of unmet menstrual health needs (e.g. products/materials, WASH infrastructure/services) on the participation and engagement of girls, women, and others who menstruate in school and work and their self-esteem and agency? | U3 |
|  | 4 | 6 | 0.919 | What are the experiences and challenges of girls, women, and others who menstruate with particular needs and circumstances (e.g. those living with HIV, those with disabilities, those who are incarcerated, those experiencing homelessness, those who have experienced FGM, trans and gender non-binary persons) in relation to their menstrual health? | U4 |
|  | 5 | 18 | 0.895 | How do financial barriers impact the ability of girls, women, and others who menstruate to manage their menstruation? | U5 |
| **Designing and implementing interventions (n=53)** | 1 | 3 | 0.93 | What new interventions could be developed to address harmful attitudes, norms and stigma and improve communication related to menstruation? | D1 |
|  | 2 | 7 | 0.915 | What characteristics of menstrual health interventions/programmes enable them to be sustained over time? | D2 |
|  | 3 | 9 | 0.913 | What new interventions could be developed to manage menstrual pain? | D3 |
|  | 4 | 10 | 0.913 | What are the impacts of unconditional and conditional cash transfer interventions on the menstrual health of girls, women, and others who menstruate, and consequently on their education, work, and social participation? | D4 |
|  | 5 | 11 | 0.913 | What are the impacts of providing free/subsidized menstrual products/materials on menstrual health? | D5 |
| **Integrating and scaling up (n=11)** | 1 | 8 | 0.915 | How can interventions to address menstrual health (e.g. education, social norm change, distribution of menstrual products/materials, improvements in WASH infrastructure/services, provision of health services for menstrual pain and disorders) be scaled up with quality and equity? | I1 |
|  | 2 | 14 | 0.902 | How can information on menstruation be integrated into existing formal and non-formal educational curricula, health services (e.g. contraceptive services, HPV vaccination, FGM support, psychosocial support), social norms, and gender equality interventions/programmes? | I2 |
|  | 3 | 15 | 0.902 | How can considerations for the menstrual health needs of girls, women, and others who menstruate with particular needs and circumstances (e.g. those with disabilities and their caregivers, those living in urban/rural settings, those who have experienced FGM, those in indigenous communities, those in humanitarian settings) be integrated into existing policies and interventions/programmes? | I3 |
|  | 4 | 17 | 0.897 | How can governments integrate menstrual health across sectors (e.g. education, health, WASH, gender) and achieve multi-sectoral coordination? | I4 |
|  | 5 | 32 | 0.876 | How can integration of menstrual health across sectors be optimally monitored and evaluated? | I5 |
| **Measurement and research (n=5)** | 1 | 1 | 0.956 | What indicators are optimal for assessing menstrual health over time (e.g. related to norms, education, health, rights, etc.)? | M1 |
|  | 2 | 19 | 0.890 | What tools/instruments/approaches/measures are optimal for assessing the impact of interventions to address menstrual health at various programmatic levels (e.g. local, national, global)? | M2 |
|  | 3 | 37 | 0.868 | What is the definition of a meaningful improvement in menstrual health, and how can it be measured? | M3 |
|  | 4 | 78 | 0.785 | How can lessons learned from the delivery of menstrual health interventions be optimally documented and shared? | M4 |
|  | 5 | 89 | 0.724 | What support is needed to encourage relevant, timely, and rigorous research on menstrual health, and which actors need to be engaged in the research process? | M5 |

**Figure 1.**
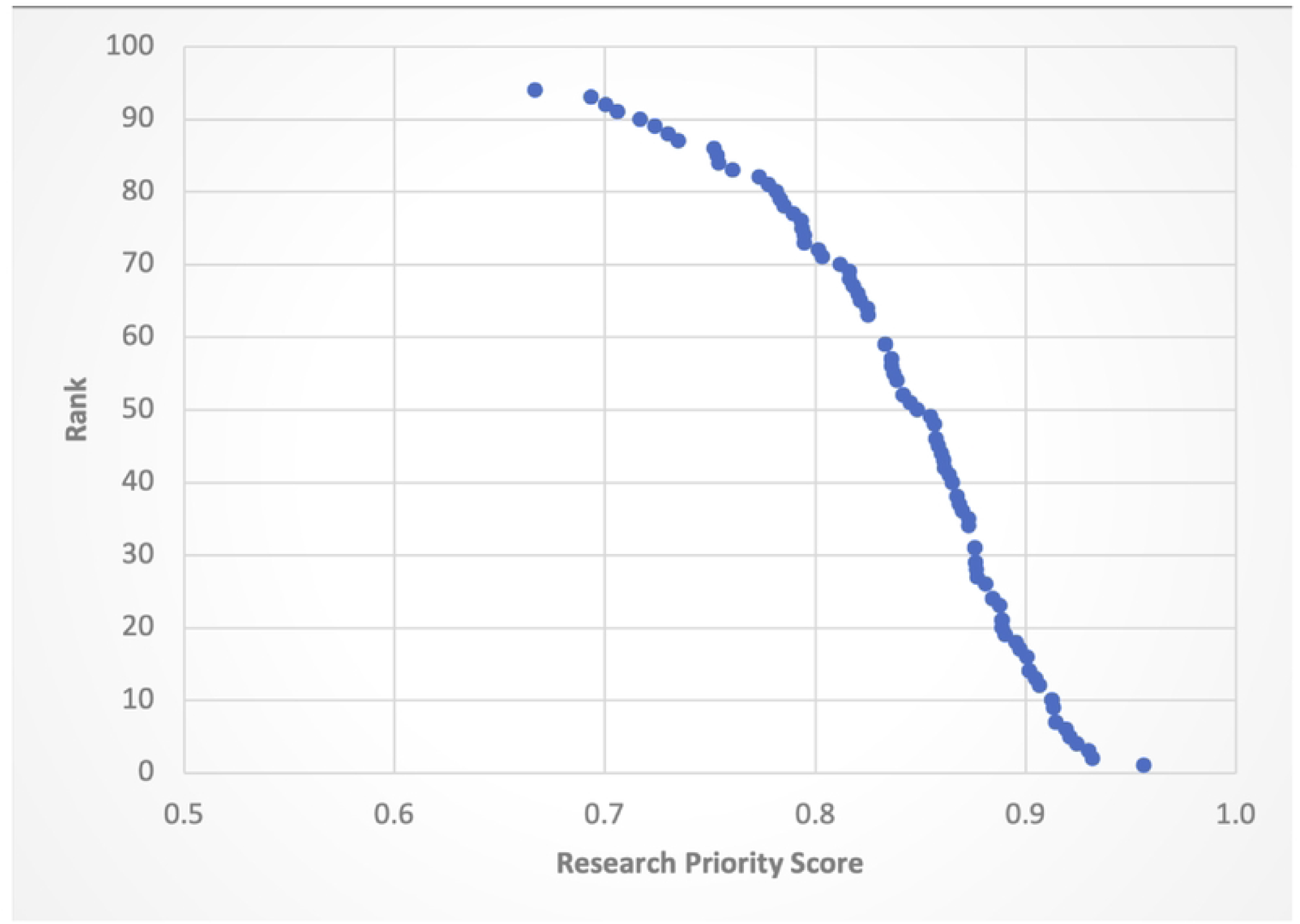
Research Priority Scores and Overall Rank.

When examining the top five-ranked research questions by individual scoring criteria, as opposed to the overall RPS, it is notable that the top five-ranked questions in the criteria ‘potential for implementation’ are in the overall top ten ranked questions by RPS, as are the top five-ranked questions in the criteria ‘importance/impact’ with the exception of one question (Table 7). In both criteria, three out of the top five-ranked questions are in the domain ‘understanding the problem.’ Additionally, it is noteworthy that the top five-ranked questions in the criteria ‘novelty’ include no questions from the domain on ‘understanding the problem’ and include instead questions from the domains on ‘designing and implementing interventions,’ ‘integration and scale-up’ and ‘measurement and research.’

**Table 7.** Top five research questions ranked by individual scoring criteria.

| Individual scoring criteria | Rank within criteria | Overall rank | Question | Overall RPS | Unique ID |
| --- | --- | --- | --- | --- | --- |
| **Novelty** | 1 | 16 | How long should menstrual cups be worn, how should they be cleaned between use, and how often should they be replaced? | 0.901 | D8 |
|  | 2 | 17 | How can governments integrate menstrual health across sectors (e.g. education, health, WASH, gender) and achieve multi-sectoral coordination? | 0.897 | I4 |
|  | 3 | 1 | What indicators are optimal for assessing menstrual health over time (e.g. related to norms, education, health, rights, etc.)? | 0.956 | M1 |
|  | 4 | 6 | What new interventions could be developed to manage menstrual pain? | 0.913 | D3 |
|  | 5 | 10 | What are the impacts of providing free/subsidized menstrual products/materials on menstrual health? | 0.913 | D5 |
| **Potential for implementation** | 1 | 1 | What indicators are optimal for assessing menstrual health over time (e.g. related to norms, education, health, rights, etc.)? | 0.956 | M1 |
|  | 2 | 4 | Are girls, women, and others who menstruate able to access and afford their preferred menstrual product/materials; what is the quality of these products/materials; where do they obtain them; and how do they use and dispose of them? | 0.924 | U2 |
|  | 3 | 3 | What new interventions could be developed to address harmful attitudes, norms and stigma and improve communication related to menstruation? | 0.930 | D1 |
|  | 4 | 2 | What are the experiences of girls, women, and others who menstruate in relation to menstrual pain and disorders (e.g. what proportion experience them, what are their perceptions about them, how do they manage them, and what support do they seek and receive for them)? | 0.932 | U1 |
|  | 5 | 6 | What are the experiences and challenges of girls, women, and others who menstruate with particular needs and circumstances (e.g. those living with HIV, those with disabilities, those who are incarcerated, those experiencing homelessness, those who have experienced FGM, trans and gender non-binary persons) in relation to their menstrual health? | 0.919 | U4 |
| **Importance/ impact** | 1 | 1 | What indicators are optimal for assessing menstrual health over time (e.g. related to norms, education, health, rights, etc.)? | 0.956 | M1 |
|  | 2 | 2 | What are the experiences of girls, women, and others who menstruate in relation to menstrual pain and disorders (e.g. what proportion experience them, what are their perceptions about them, how do they manage them, and what support do they seek and receive for them)? | 0.932 | U1 |
|  | 3 | 5 | What is the impact of unmet menstrual health needs (e.g. products/materials, WASH infrastructure/services) on the participation and engagement of girls, women, and others who menstruate in school and work and their self-esteem and agency? | 0.921 | U3 |
|  | 4 | 23 | What is the impact of unmet menstrual health needs (e.g. information, products/materials, WASH infrastructure/services) the health and wellbeing of on girls, women, and others who menstruate across the life course? | 0.888 | U6 |
|  | 5 | 3 | What new interventions could be developed to address harmful attitudes, norms and stigma and improve communication related to menstruation? | 0.930 | D1 |

When examining the average RPSs by domain, we found a similar level of prioritization of the domains among all participants, with average RPSs ranging from 0.833 to 0.862 (Table 8). However, when comparing the average RPSs by domain among academics vs. non-academics, it is notable that academics gave higher prioritization to ‘designing and implementing interventions,’ and lower prioritization to questions on ‘understanding the problem,’ ‘integrating and scaling up,’ and ‘measurement and research’.

**Table 8.** Average Research Priority Scores, by domain and stakeholder group.

| Domain | Total number of research questions | Average RPS |  |  |
| --- | --- | --- | --- | --- |
|  |  | All participants | Academics | Non-academics |
| Understanding the problem | 25 | 0.850 | 0.843 | 0.853 |
| Designing and implementing interventions | 53 | 0.833 | 0.841 | 0.830 |
| Integrating and scaling up | 11 | 0.862 | 0.850 | 0.867 |
| Measurement and research | 5 | 0.845 | 0.815 | 0.857 |

When comparing the top ten-ranked research questions among academics and those not working in academia, there are notable differences (Figure 2). First, the lists have only three questions in common (out of a total of 18 unique questions in the two lists): one regarding the optimal indicators for assessing menstrual health, one regarding interventions to address norms and attitudes about menstruation, and one regarding interventions to manage menstrual pain. Second, the top ten-ranked questions among non-academics included more questions in the domain on ‘understanding the problem’ (n=4) than those among academics (n=1). The top ten-ranked questions among academics, meanwhile, included more questions in the domain on ‘designing and implementing interventions’ (n=7) than those among non-academics (n=4).

**Figure 2.**
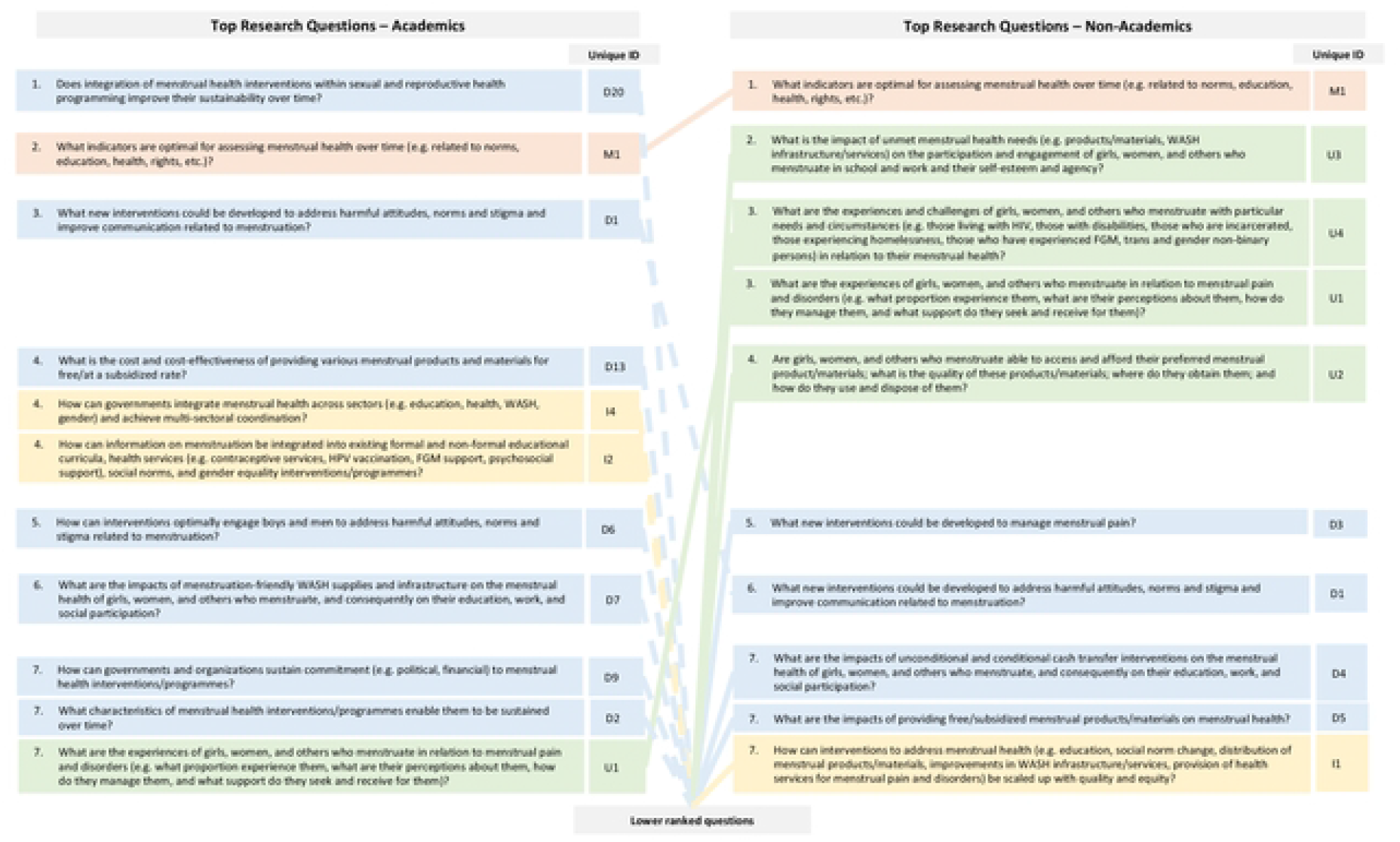
Comparison of the top ranked research questions, among academics vs. non-academic.

## Discussion

This study used a modified version of the CHNRI approach to identify research priorities on menstrual health across the life-course in LMICs, moving beyond previous research priority identification that focused on adolescent girls.^3^ In doing so, it generated input from 82 participants across all continents with expertise in policy, programming, financial support, and/or research relating to menstrual health.

Overall, the study identified a greater number of research questions in the domains of ‘understanding the problem’ and ‘designing and implementing interventions’, and higher prioritization of those research questions, than questions on ‘integration and scale-up’ and ‘measurement and research’. This confirms that there are still many knowledge gaps in understanding the menstrual experiences of women, girls, and people who menstruate, and in identifying and assessing what are the most effective and optimal interventions to meet their needs. These two domains also align with the previous research priority assessment focused on adolescent girls, which specifically noted ‘the need for a strong evidence base’, and included illustrative questions related to understanding the problem and learning around interventions.^3^ Until these gaps are addressed, it appears that stakeholders perceive questions regarding integration and scale-up to be premature. The lower prioritization of questions on ‘integration and scale-up’ may also reflect the composition of stakeholder groups that participated in this study, as only a small proportion of participants represented international agencies, government, and funding agencies.

Despite there being fewer ‘measurement and research’ questions identified and prioritized overall, the top ranked question identified, the top-ranked research question identified in this study was ‘What indicators are optimal for assessing menstrual health over time?’ The previous research priority assessment focused on adolescent girls highlighted a broad need for standardized measures, and specifically noted a need for identifying indicators for national-level monitoring for assessing changes over time. Having a standardized set of indicators is critical even to answer the other research questions in this list. A standardized set of indicators allows for comparison of menstrual health issues across and within different populations worldwide, and helps researchers, implementers, and funders to target their efforts where they are needed most. Notably, since this prioritization work was initiated, progress on indicators has been made, reflecting both the need identified in the initial research priority work with adolescent girls and the extent to which having indicators is truly wanted in the sector.^34^ Specifically, a shortlist of priority indicators for monitoring girls’ menstrual health at the national level^43^ and a list of potential indicators for monitoring menstruation among those who work outside the home have been published.^44^ Additionally, in 2021, the Joint Monitoring Programme for Water Supply, Sanitation, and Hygiene—custodians of monitoring data for SDG targets 6.1 and 6.2—included a set of harmonized menstrual health indicators as part of the first dedicated section on menstrual health in the regular reporting on household drinking water, sanitation, and hygiene.^45^

This study identified that the top research priorities were not limited to one area of expertise (e.g. education, health, WASH etc.) but were distributed across issues. For example, the top ten-ranked research priorities include questions related to menstrual pain, socio-cultural drivers of menstrual health, menstrual products, and participation in school and work. This indicates that research gaps exist in multiple domains of menstrual health, which will require addressing through collaborative efforts across all areas of expertise. Further, they identify a strong need to promote equity by understanding the specific menstrual needs and experiences of underserved populations (e.g. those living with HIV, those with disabilities, those who are incarcerated, those experiencing homelessness, those who have experienced FGM, trans and gender non-binary persons), and identifying effective interventions to meet those needs. Finally, the difference in RPS between the top 10-ranked research questions and those following was not substantial, pointing to the high prioritization of many questions beyond those included only in this abbreviated list.

This study also identified important differences in the top ten-ranked research priorities among academics and non-academics. In part, the different priorities may reflect what the two stakeholder groups see firsthand as challenges and needs in their day-to-day work. Alternatively, or perhaps in addition, it may reflect an important need for improved knowledge sharing between stakeholder groups. For example, while sorting through the proposed research questions in Phase 2, the Global Menstrual Collective’s Research and Evidence Group – which is itself largely composed of academics – felt that a number of the proposed questions already had a substantial amount of evidence in the published literature. As such, effort is needed to ensure that research evidence is not confined to academic literature, but rather that findings and recommendations are written, translated, and disseminated purposefully and meaningfully to others working in the field of menstrual health, e.g., through educational programmes, through liaison with governments to support legislation.

On the other hand, it is very possible that those directly involved in the implementation of menstrual health interventions have generated substantial learnings on interventions and how to deliver them effectively in various contexts, and thus see those domains as being of lower priority but have not had the opportunity or support to formally evaluate and/or document these learnings in the published literature. Effort is needed to ensure that these learnings are documented and integrated into the evidence base on menstrual health and disseminated across the different stakeholder groups. Finally, these differences may also reflect variations in what stakeholder groups view as worthy of the investment of what remains a limited pool of resources. For example, academics may feel that there is insufficient evidence to invest resources on an intervention, while non-academics may approach such decision-making from a different lens, such as one of human rights.

### Strengths and Limitations

This study has several limitations. First, whilst the open invitation along with snowballing technique opened our survey to a wide audience, participants were permitted to self-select and stringent criteria were not used to evaluate their eligibility for inclusion. As a result, it is possible that the study was affected by non-response bias, and some respondents may not have had sufficient expertise to be considered an ‘expert’ in menstrual health. Second, many participants began the surveys in Phases 2 and 3 but did not complete them. The format of the online survey did not allow participants to view the whole document; instead, they had to complete each page before the next page was revealed. This meant participants were unable to decide in advance whether the survey was appropriate or of interest to them until after they had completed their demographic details. A formal analysis of the barriers to completion was not possible, but we hypothesize that the length of the surveys – particularly that used in Phase 3, which had a total of 91 questions each requiring consideration of 3 criteria – may not have been user-friendly for such a wide audience. This may also have contributed to the high level of consistency in scores across the 3 criteria. Further, this was a global exercise but was only conducted in English; thus, non-English speaking menstrual health experts may not have been able to participate and, among those who did, some may not speak English as their primary language. This may be reflected in the absence of substantial participation from Latin America and the Caribbean and the Middle East. It is also important to note that the survey required participants to have stable internet connectivity, as it was not available to download; this may also have undermined full participation. As a result, the findings from this study may not perfectly reflect the opinions of all menstrual health experts. Finally, in the interest of ensuring anonymity of the data, it was not possible to confirm that the participants who responded to the survey in Phase 2 were the same as those who responded to the survey in Phase 3. As a result, the perspectives and expertise of the participants may have varied throughout the process.

However, this study also has many strengths. While research priorities were previously generated on menstrual health among school-going girls,^3^ this is the first study to generate research priorities on menstrual health across the life-course in LMICs. This is particularly timely given the growing momentum among researchers, implementers, and activists in recognizing the importance of menstrual health to female empowerment and gender equity. Additionally, the study utilized a modified CRHNI approach, which is a well-respected and widely utilized systematic approach to research priority setting with transparent criteria. The study also incorporated input from many participants representing a wide range of countries, sectors, stakeholder groups, and years of experience in menstrual health. Finally, this study included several sub-analyses to (1) understand the characteristics of those who did and did not complete the surveys and the implications of this for others seeking to use the CHNRI approach to generate research priorities, and (2) to understand how the research priorities differ by stakeholder group, along with the implications for knowledge dissemination and translation between academics and non-academics.

## Conclusions

As menstrual health continues to gain attention and emphasis as an important component of public health, it is hoped that these research priorities can be utilized by policymakers, programmers, NGOs, entrepreneurs, researchers, and funders to guide future research in this area. Recognizing that research priority setting is a dynamic process, it is also hoped that these research priorities will be revisited in an iterative manner as the field continues to evolve.

## Data Availability

All data are submitted in manuscript

## Acknowledgements

We are grateful to all members of the Global Menstrual Collective’s Research & Evidence group for their contributions to this study. Cheryl Giddings is thanked for her administrative support. We also thank the many colleagues managing collaborations and various consortia and hubs that provided a conduit to forward the surveys involved in this study to their colleagues and constituents, to ensure a wide representation of participants around the world and in different sectors. We thank all participants who so carefully generated and scored these research questions.

## Supplementary Materials

**Table S1.** Example of a consolidated research question representing similar questions.

| Consolidated Question | Individual questions listed in the survey |
| --- | --- |
| *What are the effects of female genital cutting on menstrual health?* | 'What are the effects of female genital cutting on menstrual health'? |
|  | 'How does FGM/female cutting affect a girl while menstruating and its links to infections'? |
|  | 'The impact of FGM on menstrual health (it is massive but hidden!) |

**Table S2.**
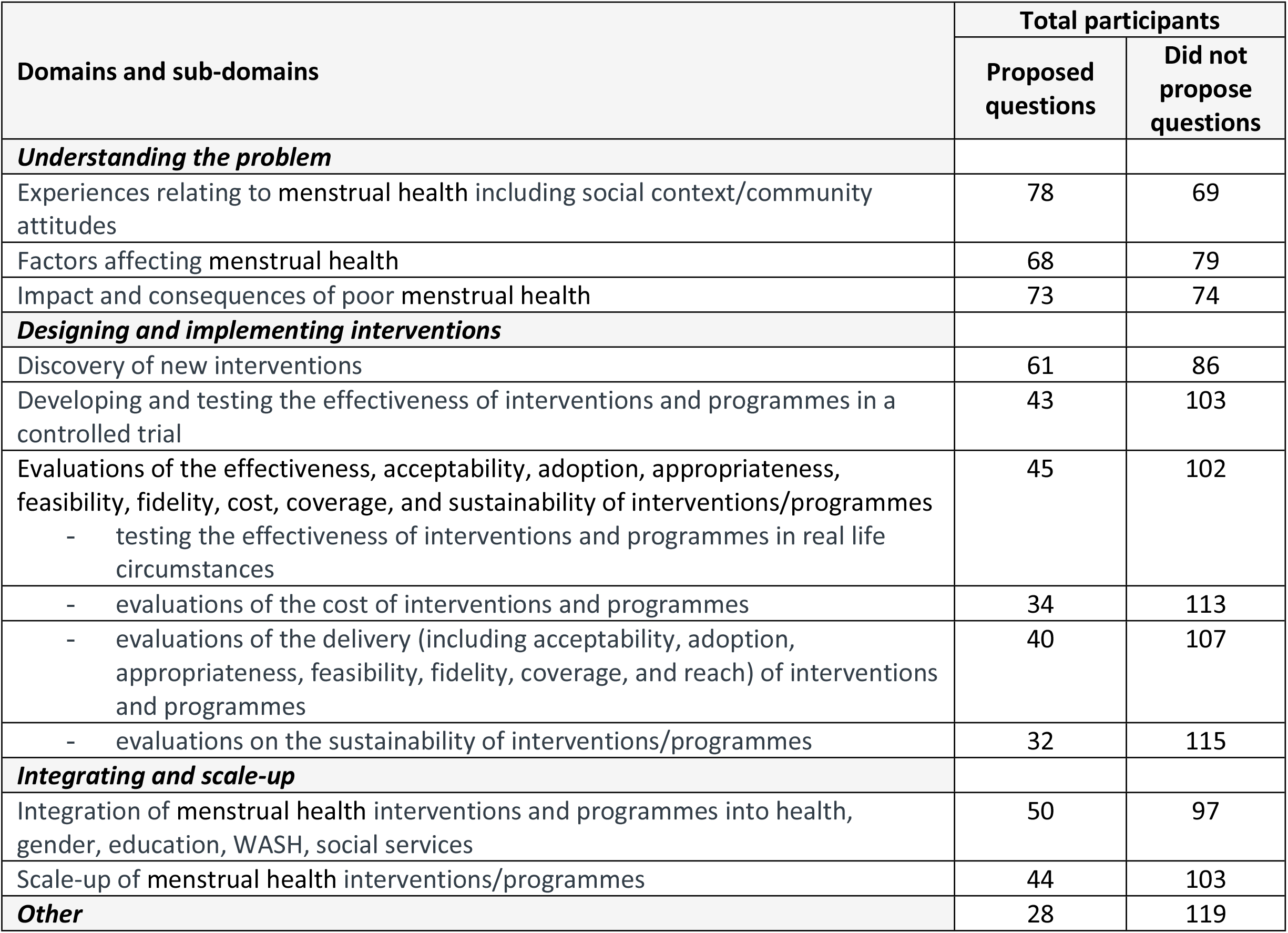
Number of participants that proposed and did not propose research questions for each sub-domain in Phase 2.

**Table S3.** Number of research questions proposed by participants that are highly experienced with 7 years or more, and less experienced (6 years or less) in menstrual health in Phase 2.

|  | Understanding the problem | Designing and implementing interventions | Integrating and scale-up | All |
| --- | --- | --- | --- | --- |
| 6 years or less | 5.33 (9.48) | 4.62 (4.42) | 1.49 (1.82) | 11.74 (7.75) |
| 7 years or more | 8.03 (7.10) | 7.10 (7.50) | 2.23 (2.14) | 17.58 (11.93) |
| all | 6.37 (3.72) | 5.57 (5.87) | 1.77 (1.94) | 13.98 (9.92) |
| p value | 0.001 | 0.065 | 0.1 | 0.009 |

